# Birth order and disease risk across the human phenome: evidence from 10 million siblings

**DOI:** 10.64898/2026.03.26.26349438

**Authors:** Benjamin Kramer, Steven A. Kushner, Andrey Rzhetsky

## Abstract

Birth order has been implicated in the etiology of individual diseases, but has never been systematically assessed at phenome-wide scale with large administrative claims data and complementary epidemiological designs. Here we use two complementary approaches: a between-family matched cohort of 1.6 million pairs and a within-family sibling comparison which includes 5.1 million families and 10.3 million individuals. These were both applied to 569 diseases defined by the ICD9-CM/ICD10-CM codes in the commercial claim data of Merative MarketScan. Of 418 diseases with adequate case counts, 150 show Bonferroni-significant birth-order associations. All odds ratios compare second-borns with first-borns, so OR *<* 1 indicates first-born excess. First-borns are at an excessive risk for neurodevelopmental conditions (autism OR = 0.74, ADHD OR = 0.93) and immune-allergic diseases consistent with the hygiene hypothesis (food allergy OR = 0.80, allergic rhinitis OR = 0.91), while second-borns are at an excessive risk for substance abuse (OR = 1.19) and gastrointestinal conditions. Between-family and within-family estimates agree in direction for 84.7% of significant diseases (Pearson *r* = 0.65), and results are robust to state fixed effects (*r* = 0.997) and full-sibling restriction. Prespecified validation controls were broadly consistent with expectations. These findings provide a comprehensive map of birth-order effects across the human disease phenome.

## Introduction

Birth order, defined as the ordinal position of a child among siblings, has fascinated researchers for over a century (Galton, 1874; Adler, 1928). Early theorists proposed that first-borns receive greater parental investment and face higher expectations, while later-borns develop in the immunological and social wake of their elder siblings (Sulloway, 1996; Ernst and Angst, 1983). These ideas have generated a rich but fragmented empirical literature, with individual studies examining birth order in relation to specific diseases or developmental outcomes.

The most influential disease-specific finding concerns allergic and atopic conditions. Strachan’s “hygiene hypothesis” proposed that younger siblings, by virtue of greater microbial exposure from older siblings during early life, develop more robust immune tolerance and lower allergy risk (Strachan, 1989). Subsequent studies confirmed protective effects of later birth order for hay fever, eczema, and asthma (Karmaus and Botezan, 2002; Ball et al., 2000; Westergaard et al., 2005; Strachan, 2000), with proposed mechanisms involving early-life microbial colonization and immune maturation (Okada et al., 2010).

A separate literature has examined birth order and neurodevelopmental conditions. Large Scandinavian registry studies demonstrated that first-borns show slightly higher educational attainment and IQ, potentially reflecting differential parental investment (Black et al., 2005, 2011; Kristensen and Bjerkedal, 2007). For autism spectrum disorder, the relationship with birth order is complicated by reproductive stoppage, or the tendency of parents to cease having children after a diagnosis. This can create artifactual first-born enrichment (Hoffmann et al., 2014; Wood et al., 2015). Interpretation is also complicated by parental-age effects, particularly the association between advanced parental age and autism risk (Durkin et al., 2008). Birth order has also been associated with psychiatric conditions and childhood mental health outcomes (Lawson and Mace, 2010), as well as metabolic diseases (Cardwell et al., 2005).

Despite this extensive literature, three limitations have impeded progress. First, nearly all studies examine a single disease or a small cluster, precluding systematic comparison of effect sizes across organ systems. Second, most designs cannot distinguish birth-order effects from confounders that covary with birth order, such as parental age, family size, socioeconomic status, and secular trends in diagnostic practice (Barclay and Myrskylä, 2016; Yang and Land, 2008). Third, sample sizes have often been too small for precise estimation, particularly for rarer conditions.

Here we address these limitations through use of a cohort of over 10 million individuals from 5.1 million two-child families in the Merative MarketScan commercial claims dataset. We employ two complementary analytic designs: a between-family matched cohort that pairs first-borns and second-borns from different families matched on sex, birth year, geography, follow-up duration, parental age, and sibling age spacing; and a within-family sibling comparison using conditional logistic regression that mitigates confounding by factors shared within families (Donovan and Susser, 2011; Frisell et al., 2012). We test 569 diseases, applying sensitivity analyses including state fixed effects and full-sibling restriction, and validate our approach with pre-specified positive and negative control diseases.

## Results

### Study design and cohort

We screened 27,975,854 enrolled families in the Merative MarketScan 2003-2024 datasets and identified 5,135,006 two-child families (10,270,012 individuals) meeting our eligibility criteria: at least one inferred parent, exactly two non-parent children, each with *≥*365 days of enrollment visibility and age at last observation *≥*12 years (Fig. 1, Table 1).

**Figure 1:**
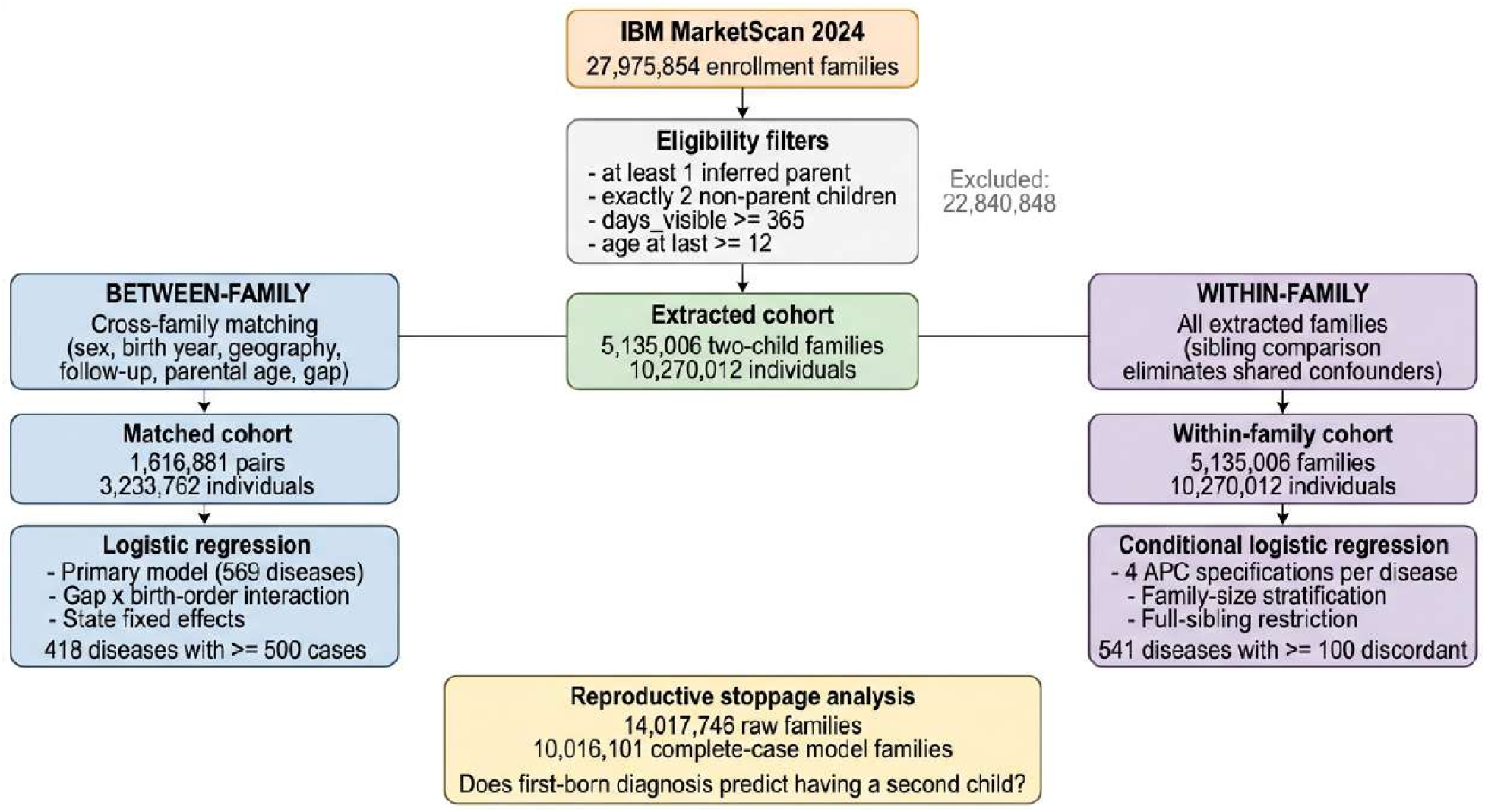
Study design and cohort flow. CONSORT-style flow diagram showing the construction of the two analytic cohorts. From 27,975,854 enrollment families in Merative MarketScan 2024, 5,135,006 two-child families (10,270,012 individuals) met eligibility cri-teria. The between-family branch created 1,616,881 matched pairs (3,233,762 individuals) via cross-family matching on sex, birth year, geography, follow-up, parental age, and sibling spacing. The within-family branch used all extracted families for conditional logistic regres-sion. A separate reproductive stoppage analysis for autism started from a raw full-database cohort of 14,017,746 families; the adjusted model used 10,016,101 complete-case families.

**Table 1:** Cohort characteristics.

| Characteristic | Value / First-born | Second-born |
| --- | --- | --- |
| *Study population* |  |  |
| Enrollment families screened | 27,975,854 |  |
| Two-child families extracted | 5,135,006 |  |
| Individuals in extraction cohort | 10,270,012 |  |
| *Between-family matched cohort* |  |  |
| Matched pairs | 1,616,881 |  |
| Individuals | 3,233,762 |  |
| Unique families | 2,164,615 |  |
| *Within-family cohort* |  |  |
| Families | 5,135,006 |  |
| Individuals | 10,270,012 |  |
| *Demographics* |  |  |
| Birth year range | 1978–2013 | 1978–2013 |
| Mean birth year | 1994.7 | 1994.7 |
| Male, $n$ (%) | 2,615,642 (50.9%) | (matched on sex) |
| Mean age at last observation (years) | 20.5 | 20.5 |
| Mean enrollment visibility (days) | 1,560 | 1,560 |
| Mean sibling age gap (years) | 3.7 | 3.7 |
| *Parental characteristics* |  |  |
| Mean paternal age at birth | 31.4 | 35.0 |
| Mean maternal age at birth | 29.4 | 32.9 |
| Father psychiatric history, $n$ (%) | 2,316,652 (22.6%) | |
| Mother psychiatric history, $n$ (%) | 3,549,504 (34.6%) | |
| *Reproductive stoppage analysis* |  |  |
| Families with $\geq 1$ eligible child (raw cohort) | 14,017,746 | |
| One-child families (stopped; raw cohort) | 4,799,163 (34.2%) |  |
| Two+ child families (continued; raw cohort) | 9,218,583 (65.8%) |  |
| First-borns with autism diagnosis (raw cohort) | 145,815 |  |
| Complete-case families in adjusted model | 10,016,101 |  |

For the between-family analysis, we identified 1,616,881 matched sibling pairs by pairing a first-born from one family with a second-born from a different family, matched exactly on sex, birth year, and urbanization tertile, with calipers on follow-up duration (*±*50 days), paternal age (*±*10 years), maternal age (*±*10 years), and sibling age gap (*±*2 years). After matching, standardized mean differences improved for all covariates. Sex, birth year, and follow-up achieved exact or near-exact balance (Supplementary Fig. 1).

For the within-family analysis, we used all 5,135,006 cohort families directly, comparing the first-born to the second-born within each family using conditional logistic regression stratified on family identifier. This design reduces confounding by factors shared between siblings (e.g., parental genetics, household socioeconomic status, geographic exposures, family health attitudes), at the cost of being powered only by disease-discordant sibling pairs (Sjölander et al., 2012).

### Phenome-wide between-family analysis

We defined 569 diseases using established ICD9-CM and ICD10-CM code groupings. Of these, 418 had *≥*500 cases in the matched cohort and were included in the between-family analysis. Logistic regression adjusted for sibling age spacing, age at last observation, sex, parental ages, parental psychiatric history, urbanization, county (via clustered standard errors), ICD coding era, enrollment time, and a full-sibling consistency flag.

Of 418 diseases tested, 150 (35.9%) reached Bonferroni significance (*P <* 1.20*×*10*^−^*^4^), and 226 (54.1%) reached significance after Benjamini–Hochberg false discovery rate correction at *q <* 0.05 (Fig. 2). Among the 150 Bonferroni-significant diseases, 79 showed first-born excess (OR *<* 1 for second-born) and 71 showed second-born excess (OR *>* 1); this deviation from a 50:50 split was not significant (exact binomial *P* = 0.568). The high rate of significant associations, together with the near-symmetric split between first-born and second-born excess, argues against a simple systematic bias inflating all associations in one direction.

**Figure 2:**
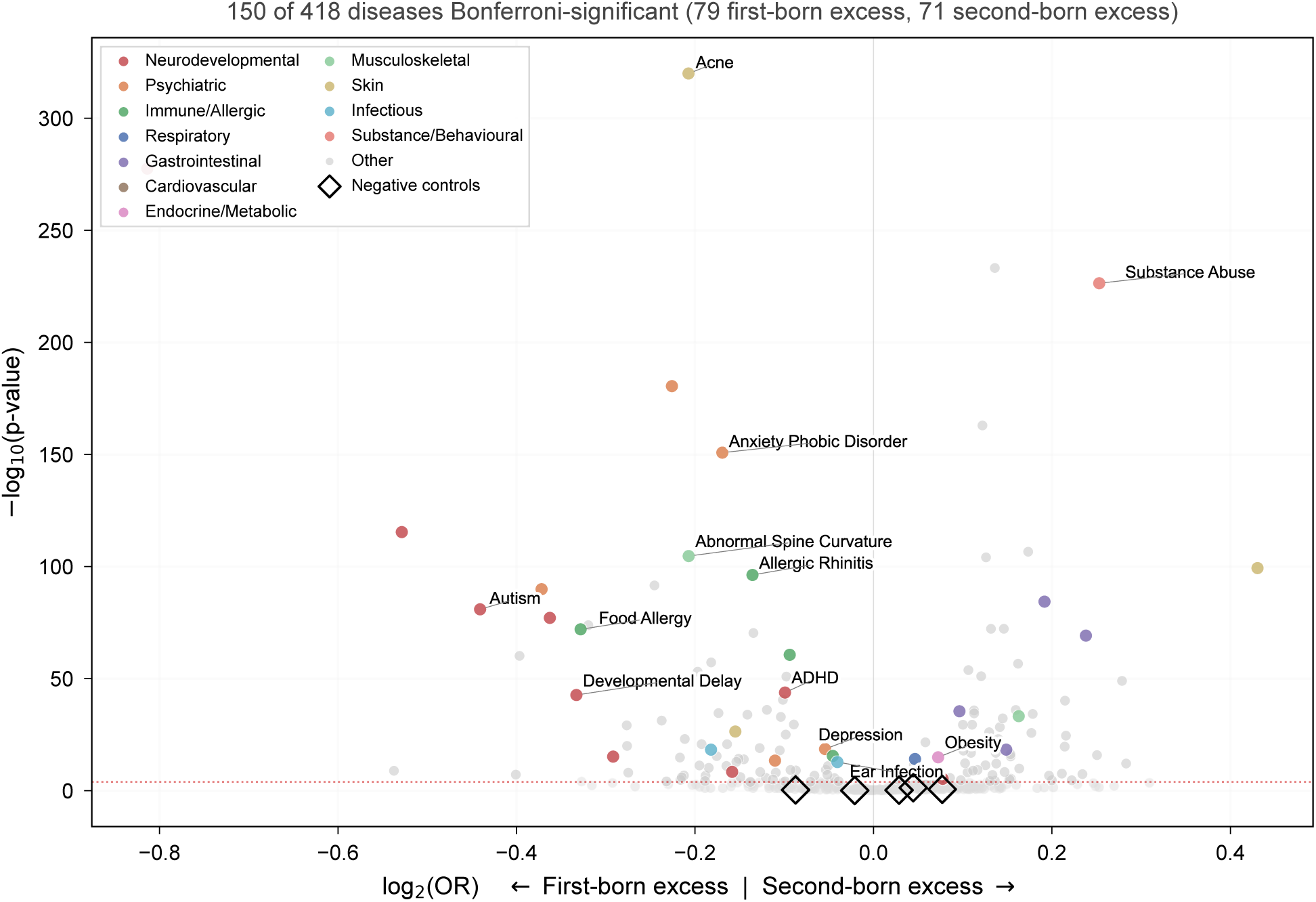
Phenome-wide birth-order associations. Volcano plot showing *−* log_10_(*P*) versus log_2_(OR) for 418 diseases in the between-family matched cohort. Points above the red dashed line are Bonferroni-significant (*P <* 1.20 *×* 10*^−^*^4^). Points with color indicate named disease categories, with gray points being non-significant. Diamond markers denote negative controls. Of 150 significant diseases, 79 show first-born excess (left) and 71 show second-born excess (right).

### Strongest birth-order associations

The 20 most significant associations (Fig. 3; Table 2) spanned multiple organ systems.

**Figure 3:**
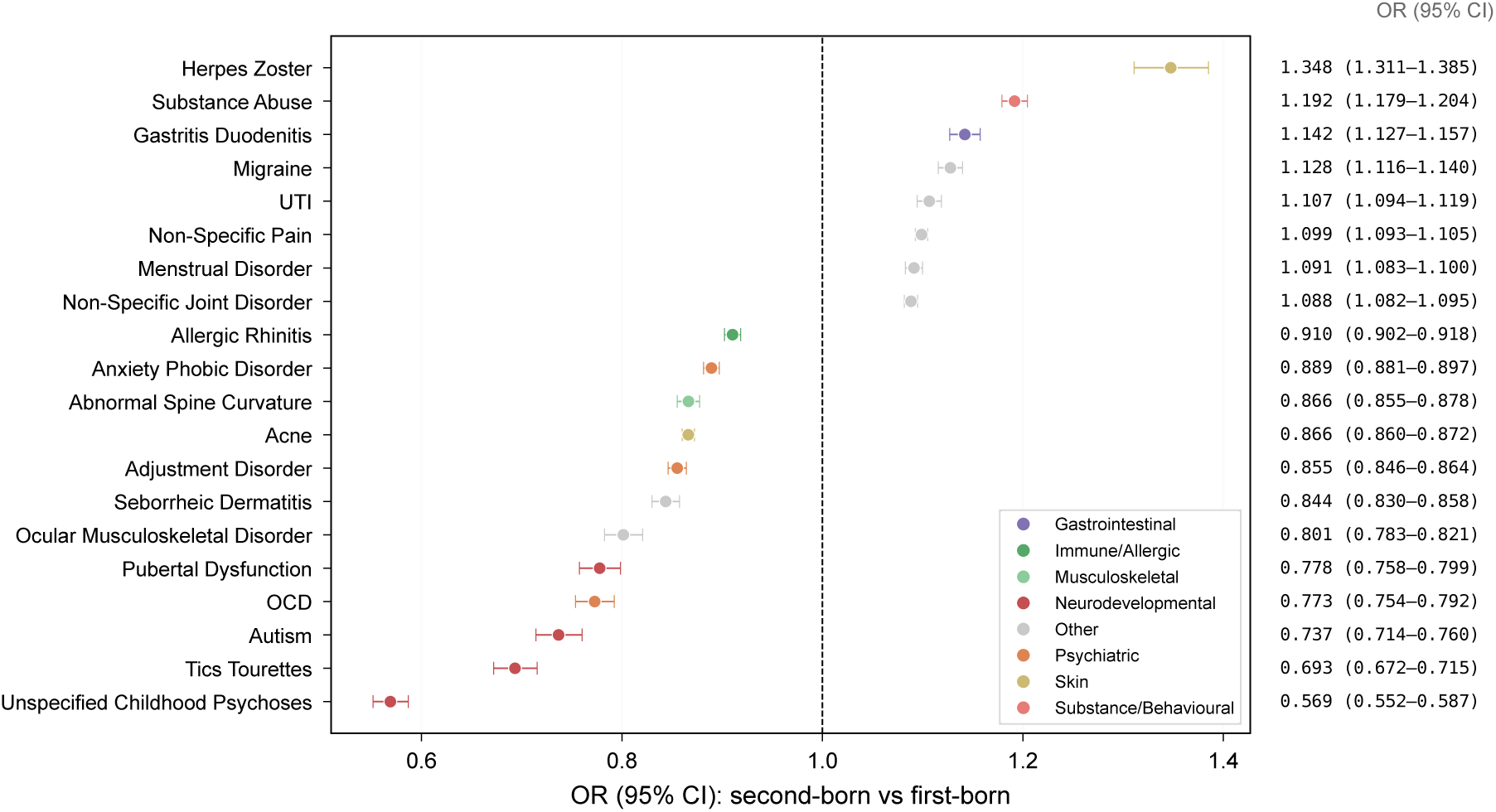
Strongest birth-order associations. A forest plot of the 20 most significant diseases, ordered by odds ratio. Points show the OR (second-born vs first-born) with 95% confidence intervals. Colors indicate disease category. Numerical OR and CI values are displayed in the right panel.

**Table 2:** Key birth-order associations across disease categories. BF = between-family matched cohort. WF = within-family sibling comparison. State FE = between-family model with state fixed effects. Agree = directional agreement between BF and WF designs. All ORs compare second-born to first-born; OR *<* 1 indicates first-born excess.

| Disease | Category | *N* | Prev. | BF OR (95% CI) | WF OR (95% CI) | FE OR | Agree |
| --- | --- | --- | --- | --- | --- | --- | --- |
| Autism | Neurodevel. | 19,985 | 0.6% | 0.737 (0.714–0.760) | 0.804 (0.786–0.823) | 0.741 | Yes |
| ADHD | Neurodevel. | 312,601 | 9.7% | 0.934 (0.925–0.943) | 0.936 (0.929–0.944) | 0.927 | Yes |
| Developmental delay | Neurodevel. | 16,943 | 0.5% | 0.794 (0.768–0.820) | 0.944 (0.911–0.978) | 0.800 | Yes |
| Anxiety/phobia | Psychiatric | 392,739 | 12.1% | 0.889 (0.881–0.897) | 0.934 (0.929–0.940) | 0.892 | Yes |
| Depression | Psychiatric | 293,022 | 9.1% | 0.963 (0.955–0.971) | 0.955 (0.948–0.962) | 0.966 | Yes |
| Bipolar disorder | Psychiatric | 58,292 | 1.8% | 0.926 (0.908–0.945) | 0.891 (0.876–0.906) | 0.931 | Yes |
| Substance abuse | Subst./Behav. | 177,202 | 5.5% | 1.192 (1.179–1.204) | 1.141 (1.129–1.153) | 1.198 | Yes |
| Allergic rhinitis | Immune/Allergic | 708,344 | 21.9% | 0.910 (0.902–0.918) | 0.911 (0.906–0.916) | 0.903 | Yes |
| Food allergy | Immune/Allergic | 36,440 | 1.1% | 0.797 (0.777–0.816) | 0.901 (0.878–0.924) | 0.810 | Yes |
| Asthma | Immune/Allergic | 399,678 | 12.4% | 0.969 (0.962–0.976) | 0.951 (0.944–0.958) | 0.976 | Yes |
| Acne | Skin | 670,949 | 20.7% | 0.866 (0.860–0.872) | 0.918 (0.912–0.924) | 0.869 | Yes |
| Spine curvature | Musculoskel. | 132,248 | 4.1% | 0.866 (0.855–0.878) | 0.927 (0.914–0.941) | 0.873 | Yes |
| Obesity | Endocrine | 112,983 | 3.5% | 1.052 (1.039–1.065) | 0.938 (0.929–0.948) | 1.058 | Flipped |
| Ear infection | Infectious | 745,900 | 23.1% | 0.972 (0.965–0.980) | 0.983 (0.977–0.989) | 0.969 | Yes |
| Type I DM | Endocrine | 18,918 | 0.6% | 1.031 (1.000–1.063) | 0.960 (0.934–0.986) | 1.030 | Flipped |
| Cystic fibrosis | Respiratory | 2,297 | 0.1% | 1.055 (0.967–1.150) | 1.127 (1.031–1.233) | 1.056 | Agree |

First-born disease risk excess was most pronounced for neurodevelopmental conditions: autism (OR = 0.737, 95% CI 0.714–0.760, *P* = 1.2 *×* 10*^−^*^81^), Tics/Tourette syndrome (OR = 0.693, 95% CI 0.672–0.715), and unspecified childhood psychoses (OR = 0.569, 95% CI 0.552–0.587). First-born excesses were also observed for acne (OR = 0.866, *P <* 10*^−^*^300^), allergic rhinitis (OR = 0.910, *P* = 5.4 *×* 10*^−^*^97^), food allergy (OR = 0.797, *P* = 9.7 *×* 10*^−^*^73^), and anxiety/phobic disorder (OR = 0.889, *P* = 1.4 *×* 10*^−^*^151^).

Second-born excess was strongest for substance abuse (OR = 1.192, *P* = 3.8 *×* 10*^−^*^227^), herpes zoster (OR = 1.348), biliary tract disease (OR = 1.179), gastritis/duodenitis (OR = 1.142), and migraine (OR = 1.128).

### Within-family sibling comparison

We used conditional logistic regression (clogit) for the analysis of the within-family cohort. We adjusted analysis for family-specific effects, for birth cohort, age at last observation in the data, sex, length of enrollment, ICD coding era, and calendar period of observation (5-year bins based on the midpoint of each child’s observation window). Of 569 diseases, 541 had *≥*100 disease-discordant sibling pairs and were analyzed further. We fitted four model specifications per disease to assess robustness to age–period–cohort parametrization including (i) cohort-adjusted, (ii) cohort plus calendar period, (iii) period only, and (iv) cohort with gap*×*birth-order interaction. The cohort-plus-period specification served as our primary within-family model.

The within-family results broadly corroborated the between-family findings. Among diseases significant in both designs, the direction and magnitude of birth-order effects were consistent: autism within-family OR = 0.804 (95% CI 0.786–0.823, *P* = 1.1 *×* 10*^−^*^74^), ADHD within-family OR = 0.936, food allergy within-family OR = 0.901, and substance abuse within-family OR = 1.141.

### Cross-design concordance

Across all diseases informative in both designs, between-family and within-family odds ratios were positively correlated (Pearson *r* = 0.648; Fig. 4). Among the 150 Bonferroni-significant diseases, 84.7% showed the same direction of effect in both designs. This concordance suggests that the observed associations are not solely driven by shared between-family confounders, because many associations persist in the within-family design, which controls for factors such as shared genetics, parental characteristics, and household environment.

**Figure 4:**
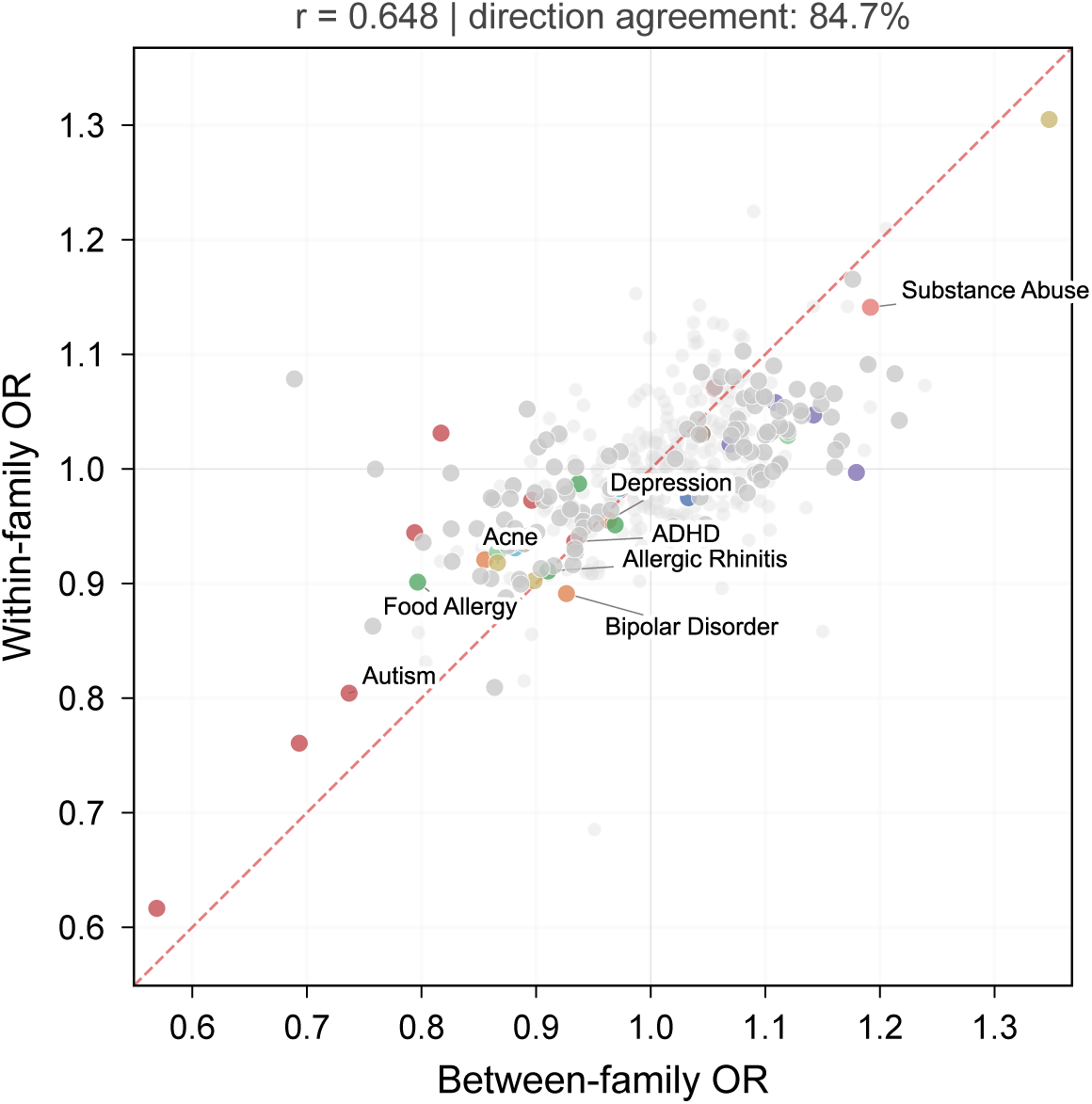
Cross-design concordance. Scatter plot comparing between-family (x-axis) and within-family (y-axis) odds ratios for diseases. The red dashed line marks perfect con-cordance (OR_BF_ = OR_WF_). Pearson for this is *r* = 0.648. Among Bonferroni-significant diseases, 84.7% show the same direction of effect in both designs.

A small number of diseases showed directional discordance between designs. The most notable was obesity (between-family OR = 1.052 indicating second-born excess, within-family OR = 0.938 indicating first-born excess). Such discordances may reflect confounders that vary within families (such as differential parental feeding practices for first vs. second children) or differential period effects on diagnosis.

### Robustness to geographic confounding

To assess whether results were driven by geographic heterogeneity in diagnostic practice, we re-estimated the between-family model with state fixed effects (50 state indicators plus the District of Columbia). The correlation between primary-model and state-fixed-effect odds ratios across all 418 diseases was *r* = 0.997 (Supplementary Fig. 2), indicating that geographic confounding has negligible influence on the birth-order estimates.

Restricting the between-family analysis to the “full-sibling” subset (families where parental-age differences are internally consistent with the sibling spacing, a heuristic for biological full siblings) produced highly concordant results (*r* = 0.998; Supplementary Fig. 4).

### Validation with positive and negative controls

We pre-specified five positive controls and seven negative controls to validate the between-family design (Fig. 5). Positive controls were diseases with established birth-order associations including allergic rhinitis, food allergy, asthma, and atopic contact dermatitis (predicted first-born excess per the hygiene hypothesis), and substance abuse (predicted second-born excess per the behavioral literature). Four of five positive controls reached Bonferroni significance in the expected direction. Atopic contact dermatitis showed the expected direction but only nominal significance (OR = 0.992, *P* = 0.041).

**Figure 5:**
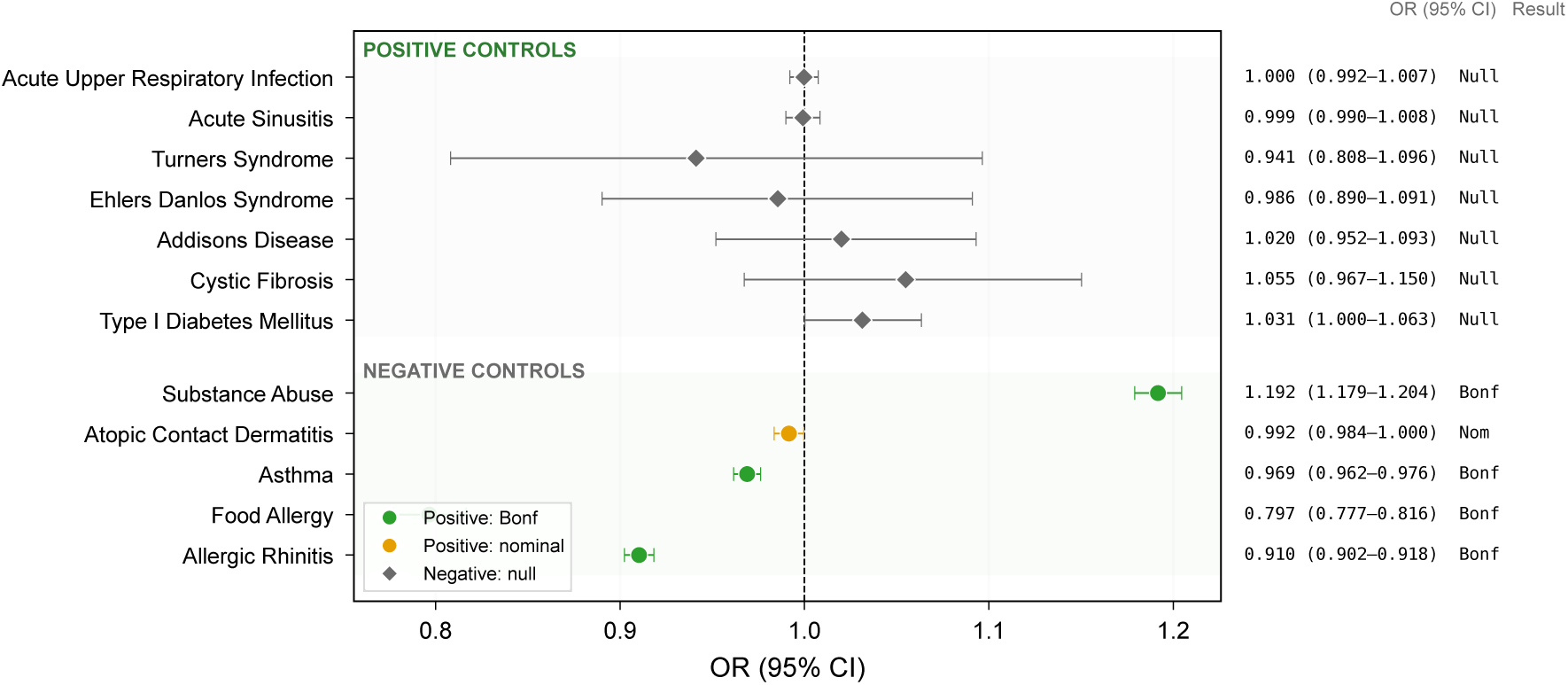
Design validation with positive and negative controls. Forest plot of pre-specified positive controls (5 diseases with established birth-order association, given the green title, gray background) and negative controls (5 diseases with primarily genetic or structural etiologies plus 2 common acute diagnoses used as empirical null comparators, gray title, green background). Four of five positive controls reach Bonferroni significance in the expected direction. Atopic contact dermatitis is directionally consistent but only nominally significant. All negative controls are non-significant at the Bonferroni threshold.

**Figure 6:**
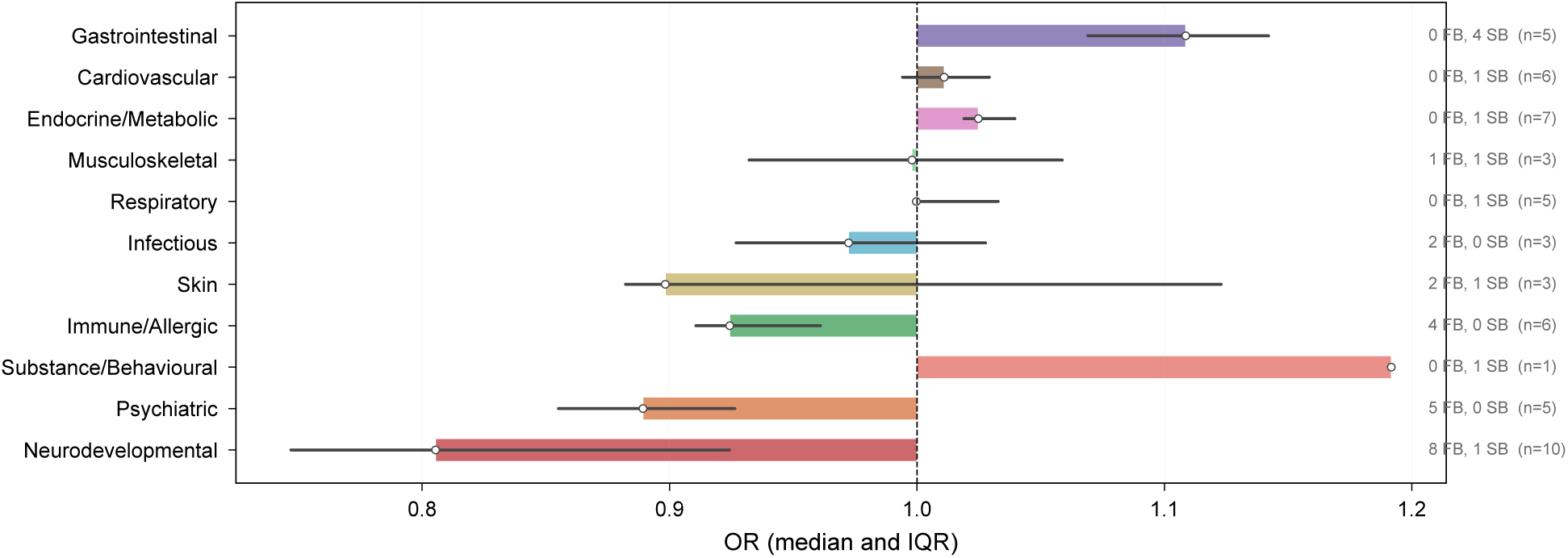
Birth-order effects by disease category. Horizontal bar chart showing the median OR and interquartile range for each disease category. The number of first-born excess (FB) and second-born excess (SB) Bonferroni-significant diseases is indicated for each category.

Negative controls included five diseases with primarily genetic or structural etiologies (type I diabetes mellitus, cystic fibrosis, Addison’s disease, Ehlers–Danlos syndrome, Turner syndrome) and two common acute diagnoses chosen as empirical null comparators (acute sinusitis and acute upper respiratory infection). All seven negative controls showed non-significant birth-order associations at the Bonferroni threshold, as expected. Corresponding within-family estimates for the same control set are reported in Supplementary Table 5.

### Sibling age spacing modulates birth-order effects

We examined whether the magnitude of birth-order effects varied with sibling age gap using a gap*×*birth-order interaction model, stratified into age gap categories (*<*4, 4–6, 7–10, *>*10 years) (Supplementary Fig. 3).

For autism, the first-born excess was strongest at gaps of 4–6 years (stratum OR *≈* 0.60) and attenuated at very short (*<*4 years) and very long (*>*10 years) gaps, producing a U-shaped pattern. Allergic rhinitis showed progressive attenuation of the first-born protective effect with increasing gap, consistent with the hygiene hypothesis (closer spacing provides more early microbial exposure from the older sibling). Substance abuse showed a decreasing second-born excess with greater spacing, suggesting that peer-influence effects of older siblings weaken when the age difference grows.

### Reproductive stoppage

To assess whether reproductive stoppage, or the tendency of parents to cease childbearing after a child is diagnosed with a serious condition, could explain the first-born enrichment observed for autism, we conducted a separate family-level analysis. The raw stoppage cohort contained 14,017,746 families with at least one eligible child in the full MarketScan database, of which 4,799,163 had only one eligible child and 9,218,583 had two or more; 145,815 first-borns had an autism diagnosis. After complete-case filtering for the adjusted model covariates, 10,016,101 families remained for inference.

A first-born autism diagnosis was associated with a modest reduction in the probability of having a second child (OR = 0.941, 95% CI 0.928–0.954, *P* = 9.4 *×* 10*^−^*^18^), corresponding to approximately a 6% relative reduction.

While statistically significant, this effect is substantially smaller than the 26% first-born excess observed in our primary autism analysis (OR = 0.737), suggesting that reproductive stoppage can account for only a fraction of the birth-order association with autism. Moreover, the within-family analysis, which is immune to stoppage bias by design, confirmed the first-born excess (within-family OR = 0.804).

## Discussion

This study provides the first phenome-wide assessment of birth-order effects on disease risk, leveraging two complementary epidemiological designs in over 10 million siblings. We identified 150 diseases with Bonferroni-significant birth-order associations spanning neurodevelopmental, psychiatric, immune-allergic, dermatological, gastrointestinal, and cardiovascular domains.

The consistency of results between designs, the performance of validation controls, and the robustness to geographic confounding collectively support the conclusion that birth order exerts statistically significant, widespread, if of modest size, effects on disease risk.

### Mechanistic interpretation

The pattern of associations suggests at least three distinct mechanistic pathways. First, the first-born excess for allergic and atopic conditions is consistent with the hygiene hypothesis (Strachan, 1989). Later-born children experience greater early-life microbial exposure from older siblings, promoting immune tolerance and reducing allergy risk. The attenuation of the allergic rhinitis effect with wider sibling spacing further supports this interpretation. Second, the first-born excess for neurodevelopmental conditions may reflect a combination of parental-age effects (first-borns have younger parents, but maternal immune priming and gestational environment differ between pregnancies), differential parental surveillance (parents may seek diagnostic evaluation more readily for their first child), and developmental programming differences between first and subsequent pregnancies (Gardener et al., 2009; Conde-Agudelo et al., 2016). Third, the second-born excess for substance abuse aligns with sociological theories of later-born risk-taking behavior and elder sibling modeling effects (Sulloway, 1996).

### Comparison with existing literature

Our findings are concordant with, and extend, prior disease-specific studies. The autism first-born excess (OR = 0.74) is consistent with prior birth-order studies but larger in magnitude, likely reflecting the combined contribution of biological birth-order effects and residual reproductive stoppage (Hoffmann et al., 2014; Turner et al., 2011). The allergic rhinitis (OR = 0.91) and asthma (OR = 0.97) effects are consistent with meta-analyses of the hygiene hypothesis (Karmaus and Botezan, 2002; Westergaard et al., 2005). The substance-abuse second-born excess (OR = 1.19) is consistent with this broader later-born behavioral framework. Our study adds hundreds of previously unexamined diseases, including strong associations for acne (OR = 0.87), adjustment disorder (OR = 0.86), biliary tract disease (OR = 1.18), and herpes zoster (OR = 1.35).

### Limitations

Several limitations merit discussion. First, claims data capture diagnoses that lead to billable healthcare encounters, not true disease incidence. Healthcare-seeking behavior may also vary with birth order. For example, if parents bring first-borns to the doctor more readily this could inflate first-born diagnosis rates independently of true disease risk. The persistence of effects in the within-family design partially mitigates this concern (within-family comparisons control for family-level healthcare-seeking tendencies), but within-family differences in birth order-dependent parental attention could remain.

Second, the age–period–cohort (APC) problem is inherent in any birth-order study. Within sibling pairs, the older child was born earlier, is observed at different ages during any calendar period, and experienced different diagnostic standards. We addressed this through cohort and period adjustment in regression, multiple model specifications in the within-family analysis, and negative controls, but residual APC confounding cannot be fully excluded.

Third, residual confounding by parental age persists despite matching and regression adjustment. The structural correlation between birth order and parental age at birth (first-borns have younger parents by definition) means that parental-age effects cannot be fully disentangled from birth-order effects without strong parametric assumptions.

Fourth, MarketScan captures employer-insured individuals, who are predominantly working-age, higher-income, and disproportionately White. Our findings may not generalize to uninsured, Medicaid-covered, or non-US populations.

Finally, the restriction to true two-child families introduces selection. Families with two children may differ systematically from larger families. This selection should be kept in mind, and future work should evaluate whether the same phenome-wide patterns replicate in larger sibships.

### Implications

These results have several implications. For clinicians, they highlight birth order as a modestly informative risk marker across multiple disease domains, which may be relevant for family counseling and screening prioritization. For researchers, the phenome-wide catalog of birth-order effects provides a resource for hypothesis generation and for benchmarking future studies. For epidemiologists, the dual-design approach demonstrated here offers a template for studying other non-randomizable familial exposures.

In conclusion, birth order is associated with disease risk more broadly than previously appreciated. The convergent evidence from between-family and within-family designs, combined with validation controls and robustness analyses, supports the conclusion that these associations reflect a mixture of biological, immunological, and social mechanisms operating across the human disease phenome.

## Methods

### Data source

We used Merative MarketScan Commercial Claims and Encounters data from the 2003–2024 annual releases, which capture inpatient, outpatient, and pharmacy claims for approximately 200 million unique covered lives in employer-sponsored health insurance plans across the United States. The raw yearly releases are distributed as SAS7BDAT files. From these annual releases, we constructed extracted analysis databases containing patient demographics (sex, birth year, enrollment family identifier), enrollment histories (start and end dates for coverage intervals), and diagnostic codes (ICD-9-CM and ICD-10-CM) recorded at each encounter (Merative, 2022).

### Cohort definition

We identified enrollment families in extracted enrollment databases derived from the annual Merative MarketScan releases, containing at least two members born between 1978 and 2013, aged 12–60 years in the current data year. Within each family, we inferred parents as the youngest male and female members who were *≥*15 years older than the oldest candidate child and whose age at the youngest child’s birth fell within 18–69 years. At least one inferred parent was required (eliminating spouse-pair misclassification).

After excluding inferred parents, we required exactly two remaining children (a “true two-child family” restriction applied before individual eligibility filters to prevent families with a third ineligible child from being misclassified as two-child families). Both children were required to have *≥*365 days of enrollment visibility, age at last observation *≥*12 years, and valid first and last observation years. The older child was designated sib order = 1 (first-born) and the younger sib order = 2 (second-born). The sibling age gap was computed as the absolute difference in birth years. Parental psychiatric history was ascertained by searching all diagnostic codes for the relevant parent against a curated set of psychiatric ICD-9/10 codes.

### Geographic annotation

Each individual was assigned a three-digit ZIP code (ZIP3) in our extracted demographics database derived from the annual Merative MarketScan files. ZIP3 was mapped to a dominant county (FIPS code) using the HUD ZIP–TRACT crosswalk, weighted by residential ratio (U.S. Department of Housing and Urban Development, 2024). County population estimates from the U.S. Census Bureau Vintage 2023 county file (co-est2023-alldata.csv) were then used to assign both Census region and county population (U.S. Census Bureau, 2024a,b). Specifically, we used the Census REGION code (1=Northeast, 2=Midwest, 3=South, 4=West) and recoded it as our analysis variable direction (E, N, S, W), where E corresponds to the Census Northeast and N corresponds to the Census Midwest. County population was also used to define the urbanization tertile (low, medium, high).

### Between-family matching

We constructed a matched cohort by pairing one first-born from family A with one second-born from family B, subject to exact match on sex, birth year, census direction, and urbanization tertile and caliper match on follow-up duration (*±*50 days), paternal age at birth (*±*10 years), maternal age at birth (*±*10 years), and sibling age gap (*±*2 years). We also had the constraint that the two individuals came from different families.

Within each exact-match stratum, we applied a greedy nearest-neighbor algorithm with randomized order and randomized tie-breaking (Rosenbaum and Rubin, 1983; Stuart, 2010). The distance metric was a weighted sum of 2.0 *×|*Δdays visible| + 1.0 *×|*Δfather age| + 1.0 *× |*Δmother age| + 0.5 *× |*Δgap|. Matching used 45 parallel workers with a fixed random seed (2025) for reproducibility.

Post-match quality control confirmed zero caliper violations, correct sib order composition for all 1,616,881 pairs, and improved standardized mean differences for all covariates (Austin, 2009) (Supplementary Fig. 1, Supplementary Table 2).

### Disease phenotyping

We defined 569 diseases using ICD-9-CM and ICD-10-CM diagnostic code groupings following previously described phenotyping approaches based on longitudinal diagnosis records (Long et al., 2023; Jia et al., 2023). A disease was considered present for an individual if any qualifying diagnostic code appeared in their claims record during the entire observation window. We imposed a minimum of 500 cases in the matched cohort (between-family analysis) and 100 discordant sibling pairs (within-family analysis) for a disease to be included.

### Between-family logistic regression

For each disease, we fitted a logistic regression model with disease status (0/1) as the outcome and birth order (sib number: 1 = first-born, 2 = second-born) as the primary exposure.

Covariates included sibling age gap (categorical: *<*4, 4–6, 7–10, *>*10 years), age at last observation (categorical), sex, paternal and maternal age at birth (categorical, 5-year bins), parental psychiatric history (father, mother), urbanization group, ICD coding era (ICD-9 vs ICD-10 based on first observation year), calendar period (5-year bins from observation mid-year), and a full-sibling consistency flag. Standard errors were clustered at the county level using HC1 robust variance (Rosenbaum and Rubin, 1983). Follow-up duration entered the model as a log-transformed covariate.

For sensitivity, we re-estimated each disease model with state fixed effects (50 state indicators plus DC).

### Within-family conditional logistic regression

For each disease, we fitted a conditional logistic regression (Cox proportional-hazards model with case–control sampling, implemented via clogit in R) stratified on family identifier, with birth order as the exposure. Covariates included birth-cohort year (centered), age at last observation (categorical), sex, log follow-up duration, ICD coding era, and calendar period of observation (categorical 5-year bins based on observation-window midpoint).

We fitted four model specifications to assess sensitivity to age–period–cohort parametrization: (i) cohort-adjusted (birth year + age + ICD era), (ii) cohort plus calendar period (primary), (iii) period only (dropping birth year), and (iv) cohort with gap*×*birth-order interaction. The cohort-plus-period specification served as the primary within-family model. Analyses were repeated in the full-sibling subset as a sensitivity analysis.

### Gap*×*birth-order interaction

In the between-family design, we estimated gap-stratified birth-order odds ratios by fitting separate logistic regressions within each gap category (*<*4, 4–6, 7–10, *>*10 years). In the within-family design, we included a gap*×*birth-order interaction term.

### Reproductive stoppage analysis

We conducted a family-level analysis using an extracted MarketScan demographics database (not restricted to two-child families). Among all families with at least one child meeting basic eligibility criteria, we identified whether the first-born had an autism diagnosis (ICD-9: 299.xx; ICD-10: F84.x) and whether the family had a second eligible child. We fitted a logistic regression with “has second child” (0/1) as the outcome and “first-born autism diagnosis” as the exposure, adjusting for first-born sex, birth year, enrollment duration, parental ages, and calendar period, with HC1 robust standard errors.

### Multiple testing correction

We applied Bonferroni correction (*α* = 0.05*/n*_tests_, where *n*_tests_ = 418 for between-family and 541 for within-family) and the Benjamini–Hochberg procedure (Benjamini and Hochberg, 1995) at FDR *q <* 0.05.

### Software

Cohort extraction and data preparation were performed in Python 3.11 (pandas, sqlite3, multiprocessing). Statistical analyses were performed in R 4.3 (survival, sandwich, lmtest packages). Figures were generated in Python using matplotlib 3.8. The analysis pipeline, figure-generation scripts, and manuscript source files are not publicly available at the time of this preprint. Public release is planned upon final publication.

### Ethics statement

This study used de-identified secondary administrative claims data from the Merative MarketScan Research Databases, which are statistically de-identified to meet HIPAA privacy requirements. The analyses used existing de-identified records, involved no direct contact with human participants, and were conducted without access to direct identifiers. On that basis, institutional review board review and informed consent were not required for this work.

### Study reporting

This preprint reports the cohort construction, disease definitions, model specifications, sensitivity analyses, and supplementary results in the manuscript and supplement.

## Data availability

Merative MarketScan data are available under license from Merative. Access requires a data use agreement. Supplementary Tables 1–5 provide summary-level results, matching diagnostics, and sensitivity analyses for the findings reported here.

## Code availability

Custom code used for extraction, matching, regression, figure generation, and manuscript assembly is not publicly available at the time of this preprint. Public release is planned upon final publication.

## Author contributions

B.K. improved the study design, implemented the analytic pipeline, performed all analyses, and wrote the manuscript.

A.R. conceived the study, supervised the study, provided critical input on study design and interpretation, designed and implemented early versions of the analytic pipeline, and edited the manuscript.

S.A.K. provided critical input on study design and interpretation and edited the manuscript.

## Data Availability

Merative MarketScan data are available under license from Merative. Access requires a data use agreement. Supplementary Tables~1--5 provide summary-level results, matching diagnostics, and sensitivity analyses for the findings reported here.

https://www.merative.com/documents/merative-marketscan-research-databases

## References

1. Francis Galton. English men of science: their nature and nurture. Macmillan, 1874.

2. Alfred Adler. Characteristics of the first, second, and third child. Children, 3:14–52, 1928.

3. Frank J. Sulloway. Born to Rebel: Birth Order, Family Dynamics, and Creative Lives. Pantheon Books, 1996.

4. Cecile Ernst and Jules Angst. Birth order: its influence on personality. Springer-Verlag, 1983.

5. David P. Strachan. Hay fever, hygiene, and household size. BMJ, 299(6710):1259–1260, 1989.

6. Wilfried Karmaus and Catalin Botezan. Does a higher number of siblings protect against the development of allergy and asthma? A review. Journal of Epidemiology and Community Health, 56(3):209–217, 2002.

7. Thomas M. Ball, Jose A. Castro-Rodriguez, Kelly A. Griffith, et al. Siblings, day-care attendance, and the risk of asthma and wheezing during childhood. New England Journal of Medicine, 343(8):538–543, 2000.

8. Tine Westergaard, Klaus Rostgaard, Jan Wohlfahrt, et al. Sibship characteristics and risk of allergic rhinitis and asthma. American Journal of Epidemiology, 162(2):125–132, 2005.

9. David P. Strachan. Family size, infection and atopy: the first decade of the “hygiene hypothesis”. Thorax, 55(Suppl 1):S2–S10, 2000.

10. Hiroshi Okada, Christoph Kuhn, Herve Feillet, and Jean-Francois Bach. The “hygiene hypothesis” for autoimmune and allergic diseases: an update. Clinical and Experimental Immunology, 160(1):1–9, 2010.

11. Sandra E. Black, Paul J. Devereux, and Kjell G. Salvanes. The more the merrier? The effect of family size and birth order on children’s education. Quarterly Journal of Economics, 120(2):669–700, 2005.

12. Sandra E. Black, Paul J. Devereux, and Kjell G. Salvanes. Older and wiser? Birth order and IQ of young men. CESifo Economic Studies, 57(1):103–120, 2011.

13. Petter Kristensen and Tor Bjerkedal. Explaining the relation between birth order and intelligence. Science, 316(5832):1717, 2007.

14. Thomas J. Hoffmann, Gayle C. Windham, Michelle Anderson, et al. Evidence of reproductive stoppage in families with autism spectrum disorder: a large, population-based cohort study. JAMA Psychiatry, 71(8):943–951, 2014.

15. Claire L. Wood, Frances Warnell, Mary Johnson, Annette Hames, Mark S. Pearce, Helen McConachie, and Jeremy R. Parr. Evidence for ASD recurrence rates and reproductive stoppage from large UK families. Autism Research, 8(1):73–81, 2015.

16. Maureen S. Durkin, Matthew J. Maenner, Craig J. Newschaffer, et al. Advanced parental age and the risk of autism spectrum disorder. American Journal of Epidemiology, 168(11): 1268–1276, 2008.

17. David W. Lawson and Ruth Mace. Siblings and childhood mental health: evidence for a later-born advantage. Social Science and Medicine, 70(12):2061–2069, 2010.

18. Chris R. Cardwell, Dennis J. Carson, and Chris C. Patterson. Parental age at delivery, birth order, birth weight and gestational age are associated with the risk of childhood type 1 diabetes: a UK regional retrospective cohort study. Diabetic Medicine, 22(2):200–206, 2005.

19. Kieron Barclay and Mikko Myrskylä. Advanced maternal age and offspring outcomes: reproductive aging and counterbalancing period trends. Population and Development Review, 42(1):69–94, 2016.

20. Yang Yang and Kenneth C. Land. Age–period–cohort analysis of repeated cross-section surveys: fixed or random effects? Sociological Methods and Research, 36(3):297–326, 2008.

21. Sharon J. Donovan and Ezra Susser. Commentary: advent of sibling designs. International Journal of Epidemiology, 40(2):345–349, 2011.

22. Thomas Frisell, Sara Öberg, Ralf Kuja-Halkola, and Arvid Sjolander. Sibling comparison designs: bias from non-shared confounders and measurement error. Epidemiology, 23(5): 713–720, 2012.

23. Arvid Sjölander, Thomas Frisell, and Sara Öberg. Causal interpretation of between-within models for twin research. Epidemiologic Methods, 1(1):217–237, 2012. doi: 10.1515/2161-962X.1015.

24. Hannah Gardener, Donna Spiegelman, and Stephen L. Buka. Prenatal risk factors for autism: comprehensive meta-analysis. British Journal of Psychiatry, 195(1):7–14, 2009.

25. Agustín Conde-Agudelo, Anyeli Rosas-Bermudez, and Matthew H. Norton. Birth spacing and risk of autism and other neurodevelopmental disabilities: a systematic review. Pediatrics, 137(5):e20153482, 2016.

26. Tychele Turner, Vasyl Pihur, and Aravinda Chakravarti. Quantifying and modeling birth order effects in autism. PLoS One, 6(10):e26418, 2011.

27. Merative. Data assets for government, non-profit, and academic research: Merative MarketScan research databases, 2022. URL https://www.merative.com/content/dam/merative/documents/brief/marketscan-data-academics-governments.pdf. Solution brief.

28. U.S. Department of Housing and Urban Development. HUD USPS ZIP Code Crosswalk Files, 2024. URL https://www.huduser.gov/portal/datasets/usps_crosswalk.html. Landing page for quarterly crosswalk releases. Accessed March 18, 2026.

29. U.S. Census Bureau. County Population Totals and Components of Change: 2020–2023, 2024a. URL https://www2.census.gov/programs-surveys/popest/datasets/2020-2023/counties/totals/co-est2023-alldata.csv. Vintage 2023 county popula-tion estimates. Accessed March 18, 2026.

30. U.S. Census Bureau. CO-EST2023-ALLDATA File Layout, 2024b. URL https://www2.census.gov/programs-surveys/popest/technical-documentation/file-layouts/2020-2023/CO-EST2023-ALLDATA.pdf. Field definitions for the county population totals file, including REGION codes. Accessed March 18, 2026.

31. Paul R. Rosenbaum and Donald B. Rubin. The central role of the propensity score in observational studies for causal effects. Biometrika, 70(1):41–55, 1983.

32. Elizabeth A. Stuart. Matching methods for causal inference: a review and a look forward. Statistical Science, 25(1):1–21, 2010.

33. Peter C. Austin. Balance diagnostics for comparing the distribution of baseline covariates between treatment groups in propensity-score matched samples. Statistics in Medicine, 28(25):3083–3107, 2009.

34. Yanan Long, Atif Khan, and Andrey Rzhetsky. Peri- and post-natal risk factors associated with health of newborns: A pregnant mother’s infections and immune diseases, and her baby’s delivery method predict immune health of the newborn. medRxiv, 2023. doi: 10.1101/2023.01.12.23284503. Preprint.

35. Gengjie Jia, Yu Li, Xue Zhong, Kanix Wang, Milton Pividori, Rabab Alomairy, et al. The high-dimensional space of human diseases built from diagnosis records and mapped to genetic loci. Nature Computational Science, 3(5):403–417, 2023. doi: 10.1038/s43588-023-00453-y.

36. Yoav Benjamini and Yosef Hochberg. Controlling the false discovery rate: a practical and powerful approach to multiple testing. Journal of the Royal Statistical Society: Series B, 57(1):289–300, 1995.

